# The exception that proves the rule: Virulence gene expression at the onset of *Plasmodium falciparum* blood stage infections

**DOI:** 10.1101/2022.12.05.22283093

**Authors:** Jan Stephan Wichers-Misterek, Ralf Krumkamp, Jana Held, Heidrun von Thien, Irene Wittmann, Yannick Daniel Höppner, Julia M. Ruge, Kara Moser, Antoine Dara, Jan Strauss, Meral Esen, Rolf Fendel, Zita Sulyok, Peter G. Kremsner, B. Kim Lee Sim, Stephen L. Hoffman, Michael F. Duffy, Thomas D. Otto, Tim-Wolf Gilberger, Joana C. Silva, Benjamin Mordmüller, Michaela Petter, Anna Bachmann

**Affiliations:** Bernhard Nocht Institute for Tropical Medicine, Bernhard-Nocht-Strasse 74, 20359 Hamburg, Germany; Centre for Structural Systems Biology, Hamburg, Germany, Notkestraße 85, 22607 Hamburg, Germany; Biology Department, University of Hamburg, Hamburg, Germany; German Center for Infection Research (DZIF), partner site Hamburg-Borstel-Lübeck-Riems, Germany; Institute of Tropical Medicine, University of Tübingen, Wilhelmstraße 27, 72074 Tübingen, Germany; German Center for Infection Research (DZIF), partner site Tübingen, Germany; Institute of Microbiology, University Hospital Erlangen, Friedrich-Alexander-University Erlangen-Nürnberg, Wasserturmstraße 3/5, 91054 Erlangen, Germany; Institute for Genome Sciences, University of Maryland, School of Medicine, 670 W. Baltimore St., Baltimore, MD 21201, USA; Cluster of Excellence: EXC 2124: Controlling Microbes to Fight Infection, Tübingen, Germany; Centre de Recherches Médicales de Lambaréné, BP242 Lambaréné, Gabon; Sanaria Inc., 9800 Medical Center Drive, Rockville, MD 20850, USA; University of Melbourne, Melbourne/Parkville VIC 3052, Australia; School of Infection & Immunity, University of Glasgow, Glasgow G12 8TA, UK

## Abstract

Controlled human malaria infections (CHMI) are a valuable tool to study parasite gene expression *in vivo* under defined conditions. In previous studies, virulence gene expression was analyzed in samples from volunteers infected with the *Plasmodium falciparum* (Pf) NF54 isolate, which is of African origin. Here, we provide an in-depth investigation of parasite virulence gene expression in malaria-naïve European volunteers undergoing CHMI with the genetically distinct Pf 7G8 clone, originating in Brazil. Differential expression of *var* genes, encoding major virulence factors of Pf, PfEMP1s, was assessed in *ex vivo* parasite samples as well as in parasites from the *in vitro* cell bank culture that was used to generate the sporozoites (SPZ) for CHMI (Sanaria® PfSPZ Challenge (7G8)). We report broad activation of mainly B-type subtelomeric located *var* genes at the onset of a 7G8 blood stage infection in naïve volunteers, mirroring the NF54 expression study and suggesting that the expression of virulence-associated genes is generally reset during transmission from the mosquito to the human host. However, in 7G8 parasites, we additionally detected a continuously expressed single C-type variant, Pf7G8_040025600, that was most highly expressed in both pre-mosquito cell bank and volunteer samples, suggesting that 7G8, unlike NF54, maintains expression of some previously expressed *var* variants during transmission. This suggests that in a new host, the parasite may preferentially express the variants that previously allowed successful infection and transmission.

## Introduction

Malaria caused by *Plasmodium falciparum* (Pf) remains one of the most serious global health problems, especially among children under the age of five. The clinical symptoms of malaria are exclusively associated with the erythrocytic stage of the parasite lifecycle, and virulence has been linked to the variant surface antigen Pf erythrocyte membrane protein 1 (PfEMP1) ^1^. This immunodominant surface antigen is encoded by approximately 60 different *var* genes that differ in the composition of their adhesive extracellular protein domains, chromosomal location, and transcriptional orientation ^2-4^. Group B *var* genes include the most telomeric genes on most of the 14 Pf chromosomes adjacent to group A *var* genes. A few group B *var* genes are also present in central chromosomal clusters along with group C *var* genes. The proteins encoded by group A are longer and more complex in domain composition than those of the other *var* gene groups ^5^. Group A and B *var* genes, particularly those containing an EPCR-binding CIDRα1 domain, are more frequently associated with severe disease and complications in paediatric infections ^6-11^. Group C *var* genes have been associated with mild malaria ^12^, and a single, highly conserved, subtelomeric, group E *var* gene encodes the VAR2CSA protein, which is strongly associated with placental malaria ^13^. A few other variants (*var1, var3*) are also conserved in various Pf isolates, but their biological function is still unknown. Although the total number of *var* genes varies among Pf isolates, the proportion within each subgroup is relatively constant ^5^. Recently, comparison of 15 genomes from geographically dispersed Pf isolates revealed that the highly polymorphic variable gene families exhibit little sequence homology, some copy number variation, but considerable consistency in their genomic organisation such as the orientation of the most telomeric *var* gene, positional conservation, and a fairly consistent number of *var* genes in internal clusters with similar orientation ^14^.

*Var* gene expression is monoallelic, i.e. each parasite expresses only a single variant at a given time ^15^. Many factors have been found to contribute to the regulation of *var* gene expression, e.g., cis-acting elements such as the *var* promoter and intron ^16,17^, trans-factors ^18,19^, higher order chromatin structures ^20,21^, and epigenetic marks ^22-26^. The majority of *var* genes is repressed in a heterochromatin environment characterized by the histone modification histone 3 lysine 9 trimethylation (H3K9me3), which is bound by heterochromatin protein 1 (HP1) ^27-29^. The single active *var* gene is largely free of H3K9me3 and instead assumes a euchromatic structure with promoter enrichment of the histone variants H2A.Z and H2B.Z and histone acetylations including H3K9ac and H3K27ac ^22,24,25,30^. Nevertheless, a comprehensive mechanistic understanding of how the mutually exclusive regulation of *var* expression occurs does not exist. Previous experiments examining switch rates in clonal cell lines showed that subtelomeric *var* genes have higher switch rates than central *var* genes, with A-type *var* genes rarely activated in *in vitro* cultures and so are not detected without selection pressure.

Mathematical modeling supports the idea of a non-random, highly structured switch pathway in which an originally dominant transcript switches either to a new dominant transcript or back to the previous one via a set of switch intermediates ^31^. The *var2csa* gene of group E was previously proposed to be one such intermediate ^32,33^. Therefore, switching of *var* gene expression is determined by intrinsic activation/deactivation rates of *var* genes, suggesting that the frequency of antigenic variation is a balanced process between hierarchical *var* switches and selective forces in the host, such as host genetics and immunity.

Controlled human malaria infections (CHMI) provide a tailored environment to study parasites *var* gene expression *in vivo* under defined conditions, e.g., variables such as host immunity and infection time (i.e., the number of replication cycles of the parasite) can be monitored or even controlled. Previous results with the Pf isolate NF54 indicate that at some point during host-to-host transmission, the *var* expression profile is reset, allowing a phenotypically diverse population of parasites expressing different subtelomeric B-type *var* genes to enter the blood ^34,35^. These results are consistent with observations in a murine malaria model, in which mosquito transmission of serially blood-passaged parasites resulted in broad activation of subtelomeric genes encoding variant surface antigens ^36^. How this reset is achieved at the molecular level is currently also unknown.

In the past, the NF54 isolate was commonly used for CHMI, but recently other strains have become available for heterologous CHMI: 7G8 from Brazil, NF166.C8 from Guinea, and NF135.C10 from Cambodia. All of these strains have been shown to be representative of their geographic origin and to differ in their genome structure, sequence, and immunogenic potential ^37,38^. The NF54 isolate was isolated from a Dutch patient who lived near Schiphol Airport, Amsterdam, and had never left the Netherlands. The infected mosquito responsible for this airport malaria case was probably imported from Africa ^39^. In contrast, 7G8 was cloned from the IMTM22 isolate in 1984 and selected for its ability to produce microgametes, exflagellate, and infect *Anopheles freeborni* resulting in oocysts and sporozoites ^40^.

In the present study we show the first *in vivo* data on *var* gene expression from CHMI with malaria-naïve volunteers infected with the Pf clone 7G8 and provide new insights into the *var* gene expression pattern that Pf uses to establish a blood stage infection after transmission from one human host to another.

## Results

### The 7G8 *var* gene repertoire and the domain composition of the encoded PfEMP1 variants

In total, the 44 *var* genes of the 7G8 clone longer than 6 kb were analyzed in this study (Table S1). An additional 24 short *var* gene pseudogenes and fragments are annotated in PlasmoDB Release 58 (23. June 2022) ^41^. To account for this, we also included a single C-type pseudogene (Pf7G8_120024200) (Table S1) ^14^.

The Pf clone 7G8 encodes six group A *var* genes, including two *var1* variants (IT- and 3D7-type ^42^), 28 group B *var* genes, ten group C *var* genes (including the Pf7G8_120024200 pseudogene) and one group E *var2csa* gene (Table S1, Figure 1A) ^14,37^. Genes of the conserved *var3* subfamily are absent. Two *var* A genes encode CIDRα1 domains that presumably can bind EPCR, and the other two A-type variants have a N-terminal head structure with CIDR8/ψ domains of unknown binding capacity. All remaining B- and C-type *var* genes encode PfEMP1 with CIDRα2-6 domains responsible for binding the host’s CD36 receptor. In total, 24 genes of type A, B or E are located in subtelomeric regions (24/44=54.6%), although it is unclear on which chromosome the *var* gene Pf7G8_000005200 is located. Three pseudogene annotations larger than 6 kB can be found on PlasmoDB: the *var1* copy (3D7-type), the group E *var2csa* gene and the B-type gene Pf7G8_060005400. All contain a premature stop codon in the ATS sequence (Table S1).

**Figure 1:**
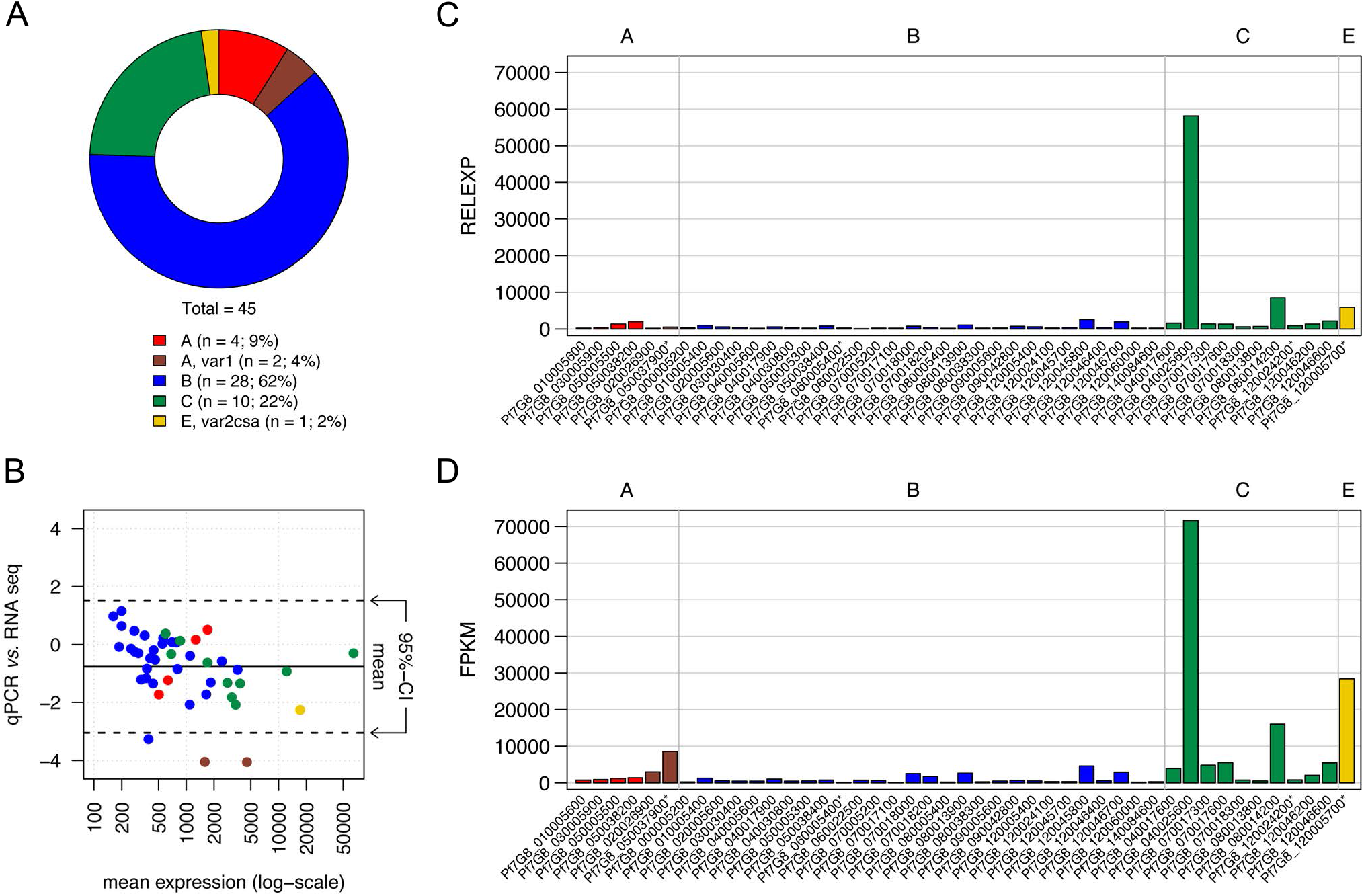
Validation of 7G8-specific qPCR covering the full *var* gene repertoire. **(A)** The genomic proportion of each *var* gene group in 7G8 parasites. **(B)** RNA-seq and qPCR expression data shown in a Bland-Altman-Plot where the mean expression of each gene is shown on the X-axis and the ratio between RNA-seq and qPCR values on the y-axis. The mean of all ratios and the confidence interval (CI) of 95% are indicated by lines. Outliers are the two *var1* genes (Pf7G8_020026900, Pf7G8_050037900) and a B-type gene (Pf7G8_060022500), which show higher expression in RNA-seq. (C, D) The RNA sample (cell bank parasites aliquot B, *in vitro* generation 13) analyzed via qPCR (C) or RNA-seq (D) shows a nearly identical expression pattern by both analysis methods. qPCR data show gene expression of each *var* gene relative to the normalizer *arginyl-tRNA synthetase*, RNA-seq data are presented in FPKM (Fragments Per Kilobase of transcripts per Million mapped reads) values. *Var* gene names are listed on the x-axis, *var* group affiliations are indicated in red (group A), dark red (group A, subfamily *var1*), blue (group B), green (group C), yellow (group E, *var2csa*).

### *In vitro var* transcript profiles of cell bank 7G8 parasites at ring stage prior to mosquito passage

Based on the newly assembled and annotated 7G8 *var* gene set, we designed and validated gene-specific primer pairs for each *var* gene (Table S2). Subsequent qPCR and RNA-seq analysis was performed side-by-side using RNA from ring-stage parasites prior to mosquito infection (Sanaria cell bank, aliquot B, generation 13). The Bland-Altman-Plot of the expression values shows high similarity between the two data sets with only three outliers, Pf7G8_020026900 (A, *var1-IT*), Pf7G8_050037900

(A, *var1-3D7*) and Pf7G8_060022500 (B-type) (Figure 1B–D).

In addition, we performed comparative qPCR analysis of two cell bank aliquots A and B to probe into putative variations within the parasite population used for sporozoite production. We showed that the *var* gene expression pattern in both aliquots is dominated by a centromeric group C *var* gene variant, Pf7G8_040025600, which accounts for 30.3% (aliquot A, generation +12 after thawing) and 43.3% (aliquot B, generation +6 after thawing) of the total *var* gene transcripts, followed by a second centromeric type C *var* gene, Pf7G8_080014200 (aliquot A: 25.2%; aliquot B: 14.9%), and the group E *var2csa* gene, Pf7G8_120005700 (aliquot A: 10.8%; aliquot B: 4.5 %) (Figure 2A, B, Table S3). This specific expression pattern was revalidated in both cell bank aliquots after additional *in vitro* proliferation cycles (aliquot A: +15, +24; aliquot B: +13, +19). It was found that the overall pattern of *var* gene expression among each aliquot remained similar over time, though Pf7G8_040025600 expression declined in aliquot A at the last time point (Figure 2A, Table S3).

**Figure 2:**
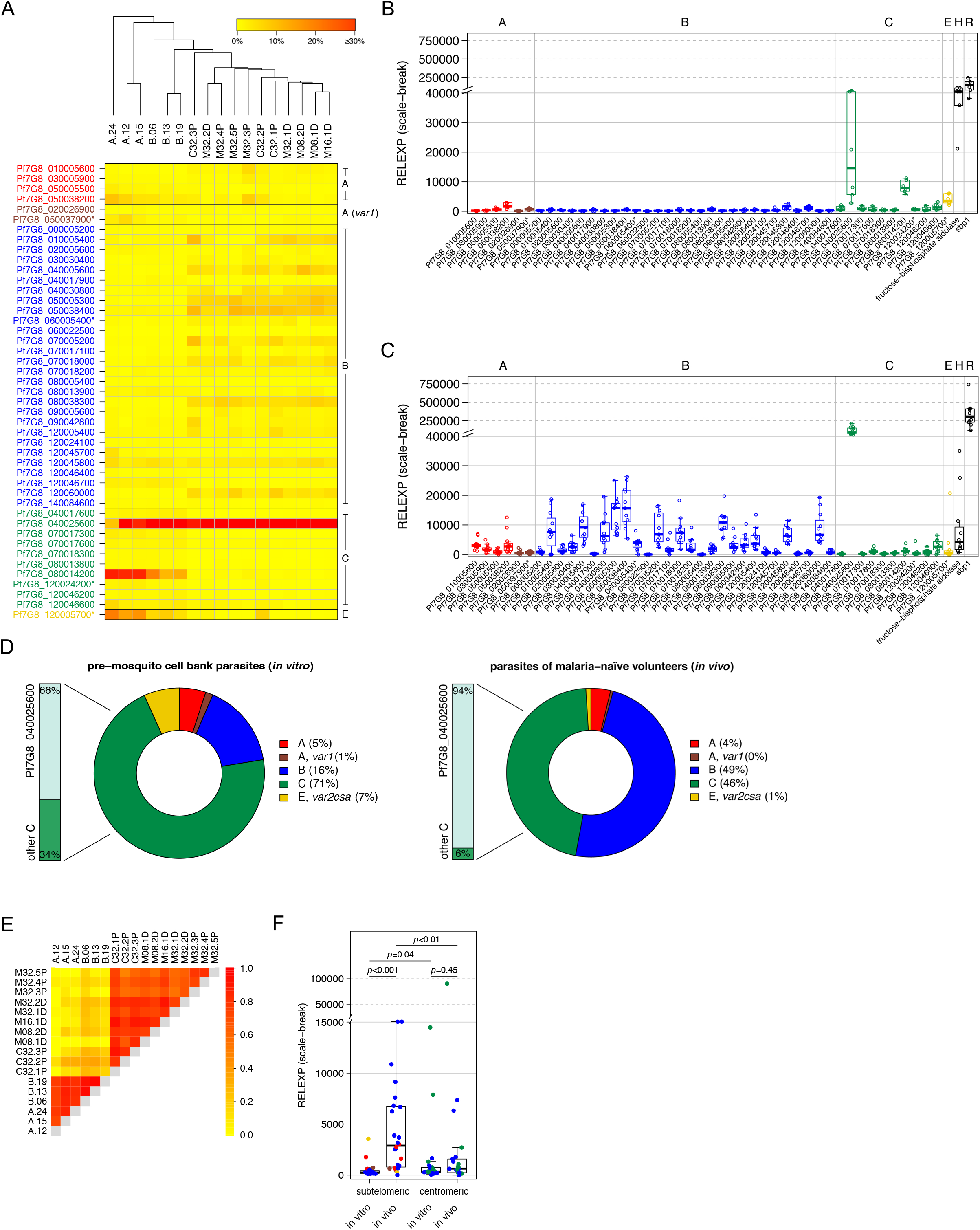
*Var* transcript profiles of pre-mosquito 7G8 cell bank parasites and of 7G8 parasites recovered from infected volunteers on the first day of detectable parasitemia. **(A)** Heat map showing individual *var* expression profiles for eleven volunteer samples (M08.1D, M08.2D, M16.1D, M32.1D, M32.2D, M32.3P, M32.4P, M32.5P, C32.1P, C32.2P and C32.3P) taken immediately before the start of treatment and six pre-mosquito cell bank parasite samples (aliquot A: *in vitro* generations 12, 15 and 24 post-thaw; aliquot B: *in vitro* generations 6, 13 and 19 post-thaw). To correct for individual differences in the total *var* expression levels, the expression for each *var* gene was normalized against total *var* expression in each sample. Hierarchical cluster analysis confirmed that pre-mosquito cell bank parasite samples differed from *in vivo* volunteer samples. (**B, C**) Gene expression of each *var* gene and controls relative to the normalizer *arginyl-tRNA synthetase* expression is shown in scatter plots for the pre-mosquito cell bank parasite line (**B)** and parasites obtained from the volunteers (**C**). Each point represents a value observed for the pre-mosquito samples taken from two independently thawed parasite stocks after 12, 15 and 24 (aliquot A) or 6, 13 and 19 parasite generations (aliquot B) and for eleven volunteer samples at day 11–13 after sporozoite inoculation. (**D**) Proportion of *var* gene expression by group for pre-mosquito cell bank parasites and parasites isolated from volunteers. **(E**) Comparison of the expression levels between subtelomeric and centromeric *var* gene variants in pre-mosquito cell bank parasites and parasites isolated from volunteers. Each dot represents the mean relative expression for each *var* gene variant located in either the subtelomeric or centromeric region of the 7G8 genome. Means and standard deviations are given for each group. Statistical analyses were performed using the Mann Whitney U test. (**F**) Heat map of pairwise Pearson correlation coefficients (PCC) between expression profiles illustrates the positive correlation between samples. Group affiliation of *var* genes is indicated by the color code with A-type *var* genes in red, the subfamily *var1* in dark red, B-type genes in blue, group C genes are colored in green and the *var2csa* gene (group E) is shown in yellow. Control genes are shown in black. Annotated pseudogenes are marked with an asterisk. H: housekeeping gene, *fructose-bisphosphate aldolase*; R: ring control, *skeleton-binding protein 1* (*sbp1*).

### *In vivo var* transcript profiles in malaria-naïve volunteers infected with 7G8 sporozoites

Next, we analyzed the *var* gene expression profiles in samples from 7G8-infected volunteers from two different clinical trials, MAVACHE ^43^ and CVac-Tü3 ^44^ (Figure S1). Our analysis of *var* gene expression profiles in parasites from eleven individual malaria-naïve volunteers (Table 1) showed that transcripts of all 45 *var* gene variants were detectable during the early blood stage 7G8 infection, similar to the expression pattern previously observed in NF54 ^35^. Interestingly, all volunteer samples showed a very similar *var* gene expression pattern (Figure 2A, C, D, E) dominated by the same single centromeric group C *var* gene variant, Pf7G8_040025600, accounting for a median 41.0% (IQR: 37.7–49.4) of total *var* gene expression (Table S3). In addition, a broad induction of subtelomeric *var* genes, mainly group B, was observed (Figure 2A, C, D). Besides the dominant Pf7G8_040025600, the next ten most highly expressed *var* variants, each accounting for more than 2% of total *var* gene expression, are classified as B-type, and nine of them are located in subtelomeric regions (Figure S2, Table S3). Hierarchical cluster analysis and pairwise Pearson correlation coefficients of samples from pre-mosquito cell bank parasites and malaria-naïve infected volunteers showed clearly distinct expression profiles except for the dominant C-type Pf7G8_040025600 highly expressed in both (Figure 2A, E). Consistent with this, a comparison of expression levels between subtelomerically located *var* genes in parasites from malaria-naïve volunteers and pre-mosquito cell bank parasites revealed a significant difference between both gene subsets, while a comparison of only centromeric *var* gene expression revealed no significant difference (Figure 2F). In general, significantly higher expression of subtelomerically located genes compared to centromeric genes was observed *in vivo*.

**Table 1:** Overview of volunteer characteristics infected with PfSPZ Challenge 7G8 and parasite counts determined either by thick blood smear (TBS) or qPCR at the day of treatment/sampling.

| Volunteer ID | Sex | Year of birth | Immunization | PfSPZ challenge | No. of sporozoites | Day of treatment | Parasites/μl (TBS) | Parasites/ml (qPCR) |
| --- | --- | --- | --- | --- | --- | --- | --- | --- |
| M08.1D | m | 1976 | dose escalation | 7G8 | 800 | 13 | 11 | 8,216 |
| M08.2D | m | 1988 | dose escalation | 7G8 | 800 | 12 | 7 | 717 |
| M16.1D | m | 1992 | dose escalation | 7G8 | 1,600 | 12 | 30 | 8,593 |
| M32.1D | m | 1989 | dose escalation | 7G8 | 3,200 | 11 | 0 | 3,671 |
| M32.2D | m | 1993 | dose escalation | 7G8 | 3,200 | 12 | 89 | 3,956 |
| M32.3P | f | 1992 | placebo | 7G8 | 3,200 | 10 | 3 | 3,560 |
| M32.4P | f | 1994 | placebo | 7G8 | 3,200 | 11 | 0 | 1,757 |
| M32.5P | m | 1991 | placebo | 7G8 | 3,200 | 11 | 0 | 3,666 |
| C32.1P | f | 1992 | placebo | 7G8 | 3,200 | 11 | 4 | 15,559 |
| C32.2P | m | 1993 | placebo | 7G8 | 3,200 | 10 | 0 | 8,955 |
| C32.3P | m | 1997 | placebo | 7G8 | 3,200 | 11 | 0 | 11,555 |

### Characterization of dominant C-type *var* gene expressing parasites

All 11 samples from 7G8-infected volunteers analyzed in this study had the same *var* expression signature: The C-type *var* gene Pf7G8_040025600 was the dominant transcript that did not appear to be reset during transmission. This might suggest that this gene in 7G8 is mis-regulated in a way that it is either i) permanently activated, or ii) escapes from the mutually exclusive epigenetic silencing machinery, or iii) not subject to resetting during transmission ^34,35^. Pf7G8_040025600 is the only *var* gene present in a central cluster of variant surface antigen genes on chromosome 4, where most other Pf strains have multiple *var* genes ^14^, and which also includes a *ruf6* sequence, a *rif* gene, and a *var* pseudogene. To probe into the first hypothesis, we re-evaluated the RNA-seq transcriptome data from the pre-mosquito cell bank parasites (aliquot B) but found no apparent activation of the other genes near Pf7G8_040025600. Some smaller non-coding transcripts were expressed at low level from upstream of this locus, including a *ruf6* sequence known to positively regulate neighboring *var* genes ^45^ (Figure 3A).

**Figure 3:**
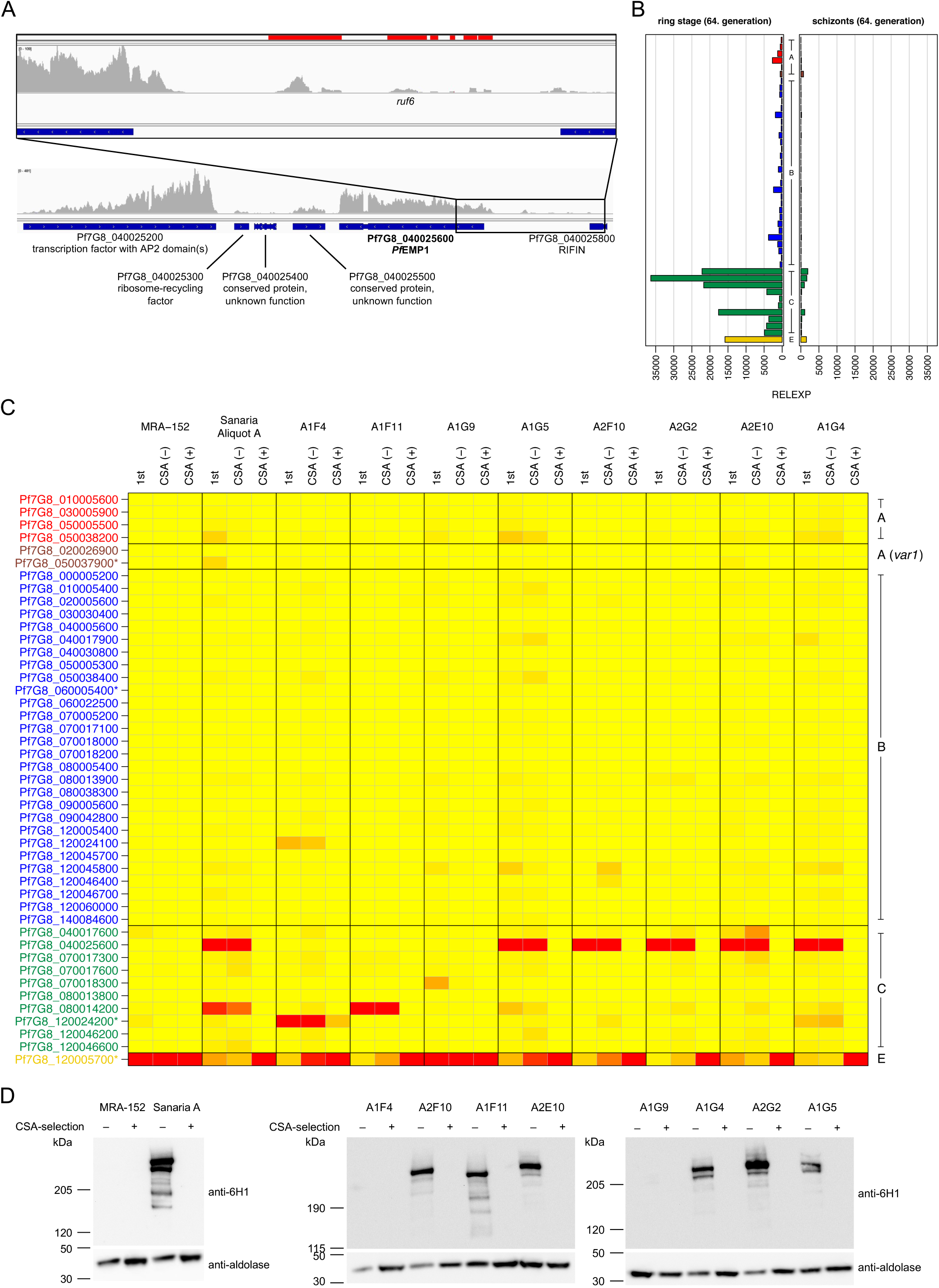
Characterization of parasites expressing Pf7G8_040025600. **(A)** Mapping of RNA-seq reads to the locus around the Pf7G8_040025600 gene in cell bank parasite aliquot B (13. *in vitro* generations after thawing). (**B**) Correct down-regulation of *var* gene expression as the parasite matures from the ring to the schizont stage (cell bank aliquot A). *Var* gene groups are indicated in the middle of the diagrams. **(C**) Selection of bulk cultures and newly generated subclones on CSA. Parasites obtained from BEI resources (MRA-152) and pre-mosquito cell bank parasites aliquot A (Sanaria A) and eight selected subclones derived from pre-mosquito cell bank aliquot A were enriched for CSA-binding. For each cell line, v*ar* gene expression is shown at three time points: (i) at the earliest time point after thawing or appearance after cloning (1^st^), (ii) before (CSA -), and (iii) after (CSA +) CSA selection. As expected from the *var* expression pattern, MRA-152 and the subclone A1G9 bound immediately to CSA due to the high expression of *var2csa* in the population. After three rounds of selection, pre-mosquito cell bank parasites and all other subclones were also enriched for CSA binding and *var2csa* expression. (**D**) Western blots of pre-selected and CSA-selected parasites using an anti-ATS antibody (6HI) and anti-aldolase as loading control. The truncation of the ATS sequence of 7G8-VAR2CSA hinders recognition of the protein by anti-ATS 6HI, explaining why PfEMP1 signal is lost in all parasite lines after CSA selection. Similarly, MRA-152, A1G9 and A1F4 show no signal due to expression of *var2csa* (MRA-152, A1G9) or the truncated pseudogene Pf7G8_120024200 (A1F4) before selection. *Var* gene names are indicated and annotated pseudogenes are marked with an asterisk, *var* gene groups are colored according to the following scheme: A in red, A-*var1* in dark red, B in blue and C in green.

Second, to ensure that Pf7G8_040025600 expression did not exhibit an aberrant expression profile during asexual blood stage development, we tightly synchronized 7G8 parasites from the cell bank and performed qPCR on ring and schizont stage parasites. The data showed that Pf7G8_040025600 expression, as well as expression of all other minor *var* transcripts, was downregulated upon parasite maturation, as expected: Schizonts showed a 22-fold reduction in relative expression compared to ring stage parasites (relative expression values of 36,357.1 in rings and 1,659.1 in schizonts) (Figure 3B).

Third, to test if Pf7G8_040025600 escaped mutually exclusive expression, we panned pre-mosquito cell bank parasites (aliquot A) on recombinant CSA, allowing selection of parasites expressing the CSA-binding PfEMP1 variant encoded by the *var2csa* gene ^46^. In parallel, an aliquot of 7G8 parasites obtained from BEI Resources (MRA-152) was subjected to the same procedure as a control for CSA binding. Immediately prior to selection on CSA, the pre-mosquito cell bank parasites expressed Pf7G8_040025600 (48.7%), Pf7G8_080014200 (17.5%) and the *var2csa* Pf7G8_120005700 at a much lower level (7.0%). In 7G8 MRA-152 parasites, the *var2csa* gene Pf7G8_120005700 clearly dominated the expression pattern even without any selection on CSA (96.2%), which was also the case in the two other 7G8 lines deposited at BEI Resources by different providers (MRA-154: 97.9%; MRA-926: 74.6%) (Figure S3). After three rounds of selection, both 7G8 lines expressed the *var2csa* gene Pf7G8_120005700 almost exclusively (cell bank aliquot A: 99.2%; MRA-152: 98.9%) (Figure 3C, D; Table S4). The conserved *var2csa* gene is known to encode the ligand for CSA ^46^, although it was annotated as a protein-coding pseudogene in 7G8 on PlasmoDB Release 58 (23. June 2022) due to a premature stop codon at position 8,622 bp resulting in a truncated ATS sequence lacking the terminal 220 amino acids (Table S1). However, in agreement with recent reports, our data suggest that the 7G8 *var2csa* gene encodes a fully functional protein that is exported to the host cell surface and can bind CSA *in vitro* ^47,48^. Fourth, another explanation for the high expression of the C-type gene Pf7G8_040025600 in pre-mosquito cell bank parasites as well as in parasites infecting the volunteers could be a very strong promoter or preference of the 7G8 strain used for CHMI to switch to Pf7G8_040025600 expression. This was investigated by cultivating the CSA-selected cell bank parasite aliquot A for an additional 100 parasite replication cycles. At every 10^th^ parasite generation, the *var* gene expression pattern was analyzed by qPCR, which was surprisingly stable over time with almost exclusive expression of the *var2csa* gene (Figure S4, Table S4). The expression of Pf7G8_040025600 was rather low, with a median relative expression value of 189.3 (IQR: 61.1–303.1) compared to the median *var2csa* expression value of 357,486.3 (IQR: 260,004.0–563,871.6). These data indicate that predominant or frequent switching to Pf7G8_040025600 expression in the 7G8 strain used is unlikely to be a major factor in upregulated expression of this gene in pre- and post-mosquito samples.

So far, all these data supported normal regulation of the Pf7G8_040025600 gene. However, because analyses at the parasite population level could miss individual variations in single parasite clones, we produced 13 clones from the cell bank aliquot A after 36 *in vitro* replication cycles by limiting dilution. The expression pattern of *var* genes was determined by qPCR, and eight clones were selected for further comparison: five of the clonal cell lines showed distinct expression of Pf7G8_040025600 (A1E4, A1G5, A2E10, A2F10, A2G2), while three cell lines dominantly expressing other *var* genes of type C (A1F4: Pf7G8_120024200, A1F11: Pf7G8_080014200) or E (A1G9: Pf7G8_120005700/*var2csa*) were chosen as controls (Figure 3C, D). These *var* expression patterns further confirm that the mechanism of mutually exclusive expression regulation in Pf7G8_040025600-expressing cell lines is still intact. Moreover, the successful selection of subclones for CSA binding revealed that *var* gene expression is not permanently fixed to this variant but can still switch to *var2csa*, which was confirmed at the RNA and protein levels (Figure 3C, D). The presence of all *var* genes on gDNA level in each subclone was verified by qPCR (Figure S5).

To identify genomic alterations in 7G8 cell bank subclones possibly associated with persistent expression of Pf7G8_040025600 after transmission, the subclones A2E10, A2G2 (both expressing Pf7G8_040025600), and A1G9 (control) were subjected to whole genome sequencing. After stringent filtering, both Pf7G8_040025600-expressing clones had mutations of A to G at position 886709 on chromosome 11, which is located in the intron of the ERO1-encoding gene Pf7G8_110027600 (putative endoplasmic reticulum oxidoreductin), and at position 679487 on chromosome 12 within the coding region of gene Pf7G8_120022100 (putative sno-RNA-associated small subunit rRNA processing protein), reflecting a synonymous exchange. Clone A2G2 had two additional single nucleotide mutations on chromosome 10 at positions 1015567 (C->T, nonsynonymous) and 1015545 (T->C, synonymous), both within Pf7G8_100029900, which encodes a conserved membrane protein of unknown function. Overall, whole genome sequencing revealed no nonsynonymous changes between the subclones expressing the Pf7G8_040025600 gene and the *var2csa*-expressing control (Table S5), making it unlikely that genetic differences between the two subpopulations cause reset failure.

Finally, to exclude epigenetic differences in gene regulation, we also performed ChIP-qPCR using ring stages from the cell bank aliquot A (bulk culture) and the same parasite line enriched for CSA-binding, as well as the cell bank subclones A2E10 (expressing Pf7G8_040025600) and A1G9 (*var2csa*-expressing control) (Figure 4, Table S6). Several epigenetic marks typically involved in *var* gene regulation were inspected, including the heterochromatin mark H3K9me3, which is associated with silent *var* genes ^22^, the euchromatic mark H3K27ac, which is typically enriched in active *var* promoters ^30^, and the histone variant H2A.Z, which occupies active *var* promoters as well as *var* introns regardless of expression status

**Figure 4:**
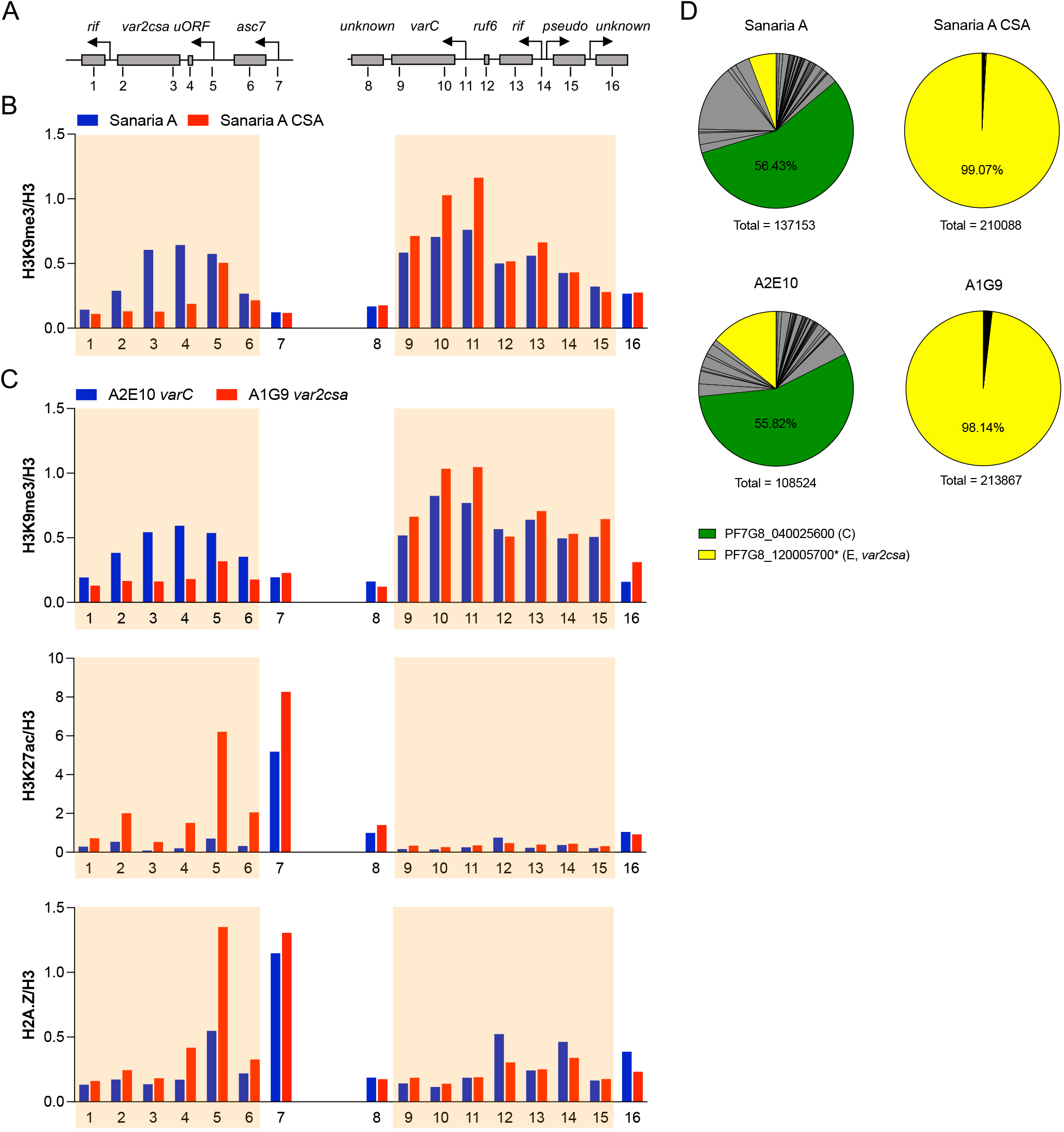
Epigenetic marks at the Pf7G8_040025600 and *var2csa* loci. **(A)** Genomic map of *var2csa* and Pf7G8_040025600 (‘*varC*’) loci with surrounding open reading frames (ORFs) and transcription start sites (indicated by arrows). The positions of the respective primer pairs used for ChIP-qPCR are indicated by numbers. (**B**) Chromatin immunoprecipitation with anti-H3K9me3 antibody and quantification of associated gDNA regions of *var2csa* (left panel) and Pf7G8_040025600 loci (right panel) by qPCR (ChIP-qPCR) in pre-mosquito cell bank parasites (aliquot A) from Sanaria (Sanaria A) and the same parasite line enriched for CSA-binding (Sanaria A CSA) (n=1). (**C**) Levels of gDNA marked with the heterochromatin-associated H3K9me3, the activation mark H3K27ac, or the alternative histone variant H2A.Z associated with active gene expression were determined by ChIP-qPCR at different regions of the *var2csa* (left panels) and Pf7G8_040025600 (right panels) loci in subclones A2E10 (expressing Pf7G8_040025600, ‘*varC*’) and A1G9 (expressing *var2csa*) (n=1). The boundaries of the predicted heterochromatin regions are shaded in B and C. (**D**) The corresponding expression profiles of *var* genes from RNA obtained in parallel with nuclei for ChIP experiments are shown as pie charts, with *var*2csa and Pf7G8_040025600 colored as indicated and all remaining *var* gene variants colored grey. Summarized total *var* gene expression is indicated for each parasite line.

^24^. Both, the CSA-enriched cell bank culture and the A1G9 subclone showed the expected epigenetic profile at their expressed *var2csa*-loci (low H3K9me3, high H3K27ac/H2A.Z) and the silenced Pf7G8_040025600 locus (high H3K9me3, low H3K27ac/H2A.Z). However, this was less obvious in unselected cell bank parasites as well as the A2E10 subclone with dominant Pf7G8_040025600 expression. The Pf7G8_040025600 gene was clearly heterochromatic at a similar level as the *var2csa* gene, and was only marginally enriched for activation marks at the promoter (H2A.Z, H3K27ac) (Figure 4B, C). However, at least a slight reduction of H3K9me3 in Pf7G8_040025600 expressing relative to *var2csa* expressing cultures was consistently observed at the Pf7G8_040025600 gene, indicating that the locus may be partially unpacked from heterochromatin in part of the population. Parallel RNA analysis revealed that selection of parasites on CSA indeed resulted in a homogenous population with very high *var2csa* expression, whereas the unselected cell bank parasites and the A2E10 subclone showed a more diverse pattern of *var* expression with lower levels of total *var* expression and Pf7G8_040025600 in particular, which may possibly explain the differences in signal intensity of the epigenetic marks (Figure 4D).

Overall, we demonstrated that expression of the C-type gene Pf7G8_040025600 (i) is a unique feature of parasites from the Sanaria cell bank, (ii) is correctly regulated across asexual blood stages, (iii) occurs in a mutually exclusive manner, and (iv) parasites are neither fixed to nor tend to preferentially activate expression of this variant after selection for *var2csa* expression. Moreover, parasites expressing this *var* variant share the genomic and epigenetic background with parasites expressing other *var* variants, which further rules out the presence of two distinct 7G8 parasite populations in the original cell bank aliquots, with one population behaving like NF54 and resetting *var* gene expression during mosquito passage, and the other population having lost its ability to reset *var* expression and being fixed to express Pf7G8_040025600. Therefore, our data rather support the second hypothesis, namely that Pf7G8_040025600 is not subject to resetting during transmission, suggesting that epigenetic resetting of *var* genes can be incomplete during passage through mosquitoes, possibly through the semiconservative retention of histone modifications at replicated *var* loci during meiosis.

## Discussion

After transmission, parasites are faced with a new environment defined by various parameters such as the metabolic state of the host, pre-existing immunity, drug pressure, pregnancy or other infections. One way to cope with these alterations might be to epigenetically reset genes upon transmission to create a diverse population of parasites expressing different *var* genes from which clones best adapted to the new environment can expand. This “bet-hedging” strategy, which is a viable alternative to directed transcriptional responses, is thought to be key to adaptations to general variations in the environment ^49^. This has been shown, for example, for expression of *var* genes after liver release of NF54 parasites in CHMI of immunologically naïve individuals ^34,35,50,51^. There, expression of *var* genes at the onset of infection shows a broad activation pattern of many group B subtelomeric genes and certain group A variants. This broad expression repertoire is reduced in NF54 parasites from infected volunteers with a higher degree of pre-existing immunity. In these individuals, only a single or very few *var* gene variants are highly expressed, presumably encoding PfEMP1 which are not recognized by the host immune system ^33^. Intriguingly, the *var* gene expression profile changed dramatically during mosquito passage of NF54 Pf parasites from the monoallelic *var2csa* expression in the initial culture used for gametocytogenesis and sporozoite production to the broad pattern of subtelomeric *var* gene transcription in the infected volunteers ^35,51,52^.

In this study, we investigated whether the observations using NF54 parasites also hold true in other Pf isolates, or whether parasite strain-specific differences in *var* gene expression patterns can be observed. This is of particular importance because current vaccination concepts using whole parasites – either radiation-attenuated sporozoites (PfSPZ Vaccine) or sporozoites co-administered with chemoprophylaxis (PfSPZ-CVac) – are based on the strain NF54. For the first time, we have obtained expression data for the entire *var* gene repertoire in another parasite strain, 7G8, which originates from a different geographic region and is genetically distinct from NF54 ^37,38^. When 7G8 parasites were used for CHMI, a very similar pattern of *var* gene group B activation was observed in naïve volunteers. This suggests that Pf generally preferentially activates subtelomeric *var* genes after release from the liver, while central *var* genes tend to remain silent ^35^, allowing a phenotypically diverse population of parasites expressing different subtelomeric *var* genes to enter the blood from the liver. These results are consistent with observations in a mouse malaria model, where transmission of serially blood-passaged parasites by mosquitoes resulted in broad activation of subtelomeric genes encoding variant surface antigens ^36^. How this switch occurs at the molecular level is currently unknown, but there is strong evidence that the regulation of *var* genes in Pf relies on epigenetic mechanisms ^52-54^.

The broad activation pattern of many *var* genes seems to contradict the concept of safeguarding the antigenic repertoire. However, this strategy of releasing a parasite population from the liver that expresses PfEMP1 proteins with diverse adhesive properties would allow optimal and rapid adaptation to a new host environment shaped by selective forces such as host genetics and pre-existing immunity. At the same time, maintaining a repertoire of *var* gene variants silenced after liver release would prevent the parasites from exposing their entire antigenic repertoire too early. Differences in intrinsic switching rates of individual *var* genes would then lead to the expansion of parasite populations that express dominant PfEMP1 variants during the course of infection and are further selected by immunity, consistent with the fashion observed in chronic infections.

Interestingly, the dominant C-type *var* gene Pf7G8_040025600 in our study escapes from resetting during transmission. There are at least five possible explanations for the high expression of this variant in pre-mosquito parasites and *in vivo* during infection of volunteers: (i) the gene might have escaped mutually exclusive expression, (ii) Pf7G8_040025600 might be generally deregulated during asexual blood stage development, (iii) a subpopulation of parasites might have a mutation or carry epigenetic marks making this variant resistant to the resetting mechanism during mosquito passage, (iv) this *var* gene variant could have a very active promoter that allows high expression levels under different conditions, or (v) only parasites expressing *var2csa* or other *var* gene variants that act as switching hubs are able to reset their *var* gene expression during transmission. We tested most of these hypotheses and demonstrated that Pf7G8_040025600 expression is mutually exclusive with that of other *var* genes, and the gene is correctly repressed in schizonts, does not have a higher on switch rate than other *var* gene variants, and none of the few genetic differences observed between clonal Pf7G8_040025600- and *var2csa*-expressing parasites can explain the phenotype. Moreover, these parasites were still able to switch their *var* expression to a different variant, consistent with the successful generation of 7G8 sporozoites by Sanaria, suggesting that the switch from asexual replication to gametocytogenesis is also not hindered in these parasites. Our results from ChIP-qPCR also suggest that epigenetic regulation of Pf7G8_040025600 is not affected, as the gene is normally silenced by heterochromatin. The absence of detection of activating histone marks may be explained by the mixed *var* expression pattern in about half of the population, as it is known that in parasite populations with heterogeneous *var* gene expression, signals from ChIP-qPCR or -seq can be leveled out ^51,55^.

Based on our data, we hypothesize that *var* gene expression may partly underlie epigenetic imprinting during transmission from one human host to another, which is thought to depend on the ability of each *var* gene locus to establish and maintain heterochromatin. Consistent with other studies showing that C-type *var* genes are turned on less frequently but can dominate expression patterns by slow silencing ^56,57^, C-type *var* genes may be less effective at re-establishing heterochromatin during transmission, in line with their location in central chromosomal regions with likely fewer Sir2A-dependent silencing nucleation sequences. In conjunction with the intrinsic promoter activity of each *var* gene variant, Pf could exhibit a loose activation hierarchy of subtelomeric *var* genes upon entry into the human blood phase. This strategy would allow the parasite population to explore a new host environment by expressing many different PfEMP1 variants as well as testing the previously successful variant in a different host, adding another layer to the parasite’s survival or adaptive strategies in humans. Although purely speculative, this could also explain why some parasite strains cause more severe disease courses than others. However, since *var2csa* is also discussed as a switching intermediate ^31-33^, it is possible that resetting is also coordinated via intermediate *var2csa* expression, from which parasites can express *var* genes at the onset of the blood phase with probabilities based on intrinsic on-rates for each variant. It should be noted that truncation of VAR2CSA-ATS in 7G8 parasites could also affect the efficiency of these processes. Further studies, such as CHMIs with NF54 parasites or other strains expressing different *var* gene variants prior to transmission, or with parasites with genetically modified *var2csa* locus, are needed to evaluate these hypotheses.

In summary, we demonstrated that (i) expression of subtelomeric B-type *var* genes is induced in 7G8 parasites at the onset of blood stage infection in malaria-naïve individuals, (ii) cell bank parasites used for PfSPZ production and isolated from volunteers exhibit an expression pattern dominated by a single C-type variant, Pf7G8_040025600, suggesting that this C-type variant underlies epigenetic memory during mosquito passage. Our results from two genetically distant parasite backgrounds show that expression of virulence-associated genes in Pf is, at least partially, reset to subtelomeric B-type expression during transmission from mosquito to human host, but also provide evidence for an alternative strategy of the parasite in which infection in the next host is established by maintenance of expression of a previously successful PfEMP1 variant in part of the parasite population. This suggests that the PfEMP1 variant expressed in the previous malaria patient is an important factor that could determine the pathophysiology of the subsequent infection. In conclusion, the NF54 strain and its clone 3D7 appear to be an accurate reference for the entire species in terms of gene content and organization, as previously noted by Otto et al. ^14^, but there appear to be differences among Pf isolates in resetting *var* gene expression during transmission to establish infection in another human host ^34,35^.

## Methods

### Ethics

The ethics committee of the University Clinic and the Medical Faculty of the University of Tübingen approved both studies, MAVACHE and CVac-Tü3, of which samples were examines in this work, and the U.S. Food and Drug Administration Agency (FDA) provided regulatory oversight. The studies were conducted according to the principles of the Declaration of Helsinki in its 6th revision and the guidelines of the International Conference on Harmonization–Good Clinical Practice (ICH-GCP). For the MAVACHE study, the registration code at ClinicalTrials.gov is NCT02704533; the CVac-Tü3 trial is registered with the EU Clinical Trial Register under 2018-004523-36. All volunteers provided written informed consent, and understanding of the study and procedures was assessed with a quiz.

#### CHMI trials and blood sampling

Samples were collected during different phases of the MAVACHE ^43^ and CVac-Tü3 trials ^44^. Both trials were conducted at the Institute of Tropical Medicine in Tübingen, Germany, where healthy, malaria-naïve volunteers were infected with live, infectious, aseptic, purified, cryopreserved NF54 or 7G8 sporozoites (Sanaria® PfSPZ Challenge (NF54) and Sanaria® PfSPZ Challenge (7G8)) manufactured by Sanaria Inc., USA, 7G8 under license from Walter Reed Army Institute of Research. On the day of treatment, up to 50 mL of blood was taken from all volunteers into sodium citrate tubes and processed by Ficoll gradient centrifugation followed by filtration of the washed red blood cell pellet through a Plasmodipur filter (EuroProxima).

The MAVACHE trial aimed to sequentially optimize the dose and schedule of PfSPZ Vaccine, verified by randomized, controlled, double-blind immunization and controlled human malaria infection in malaria-naïve, healthy adult volunteers in Germany. The dose optimization phase included a dose-finding phase to evaluate the safety, tolerability and infectivity of 7G8 PfSPZ in malaria-naïve, healthy adult volunteers. A total of nine volunteers received either 800 (n=3), 1,600 (n=3) or 3,200 (n=3) 7G8 PfSPZ. Similar to NF54, 7G8 PfSPZ at a dose of 3,200 PfSPZ resulted in parasitemia in 3/3 of the volunteers, and 2/3 volunteers developed parasitemia after infection with 800 and 1,600 PfSPZ, respectively, all administered by direct venous inoculation. Parasite kinetics and clinical presentation are similar to CHMI with NF54 PfSPZ ^58^. Samples from five volunteers are included in our study (800 PfSPZ: M08.1D, M08.2D; 1,600 PfSPZ: M16.1D; 3,200 PfSPZ: M32.1D, M32.2D) (Table 1, Figure S1). During the regimen verification phase, volunteers were either infected with PfSPZ Vaccine or received placebo, followed by a CHMI three weeks after the last immunization. The placebo group received normal saline instead of PfSPZ Vaccine, but also underwent subsequent CHMI. Samples from three volunteers infected with 7G8 PfSPZ (M32.3P, M32.4P, M32.5P) were included in our study (Table 1, Figure S1).

The CVac-Tü3 trial assessed the safety and protective efficacy of a simplified Pf sporozoite Chemoprophylaxis Vaccine (PfSPZ-CVac) regimen in healthy malaria-naïve adults in Germany ^44^. In total, NF54 PfSPZ of PfSPZ-CVac was administered three times (day 0, 5 and 28) to volunteers receiving parallel chloroquine treatment (1.1 × 10^5^ PfSPZ each), and these volunteers underwent CHMI with 3,200 7G8 PfSPZ ten weeks after the last immunization. Samples from three placebo-infected volunteers (C32.1P, C32.2P, C32.3P) could be included into our study (Table 1, Figure S1).

In total, samples from eleven malaria-naive volunteers infected with 7G8 PfSPZ were included in this study.

### Parasite cell culture

7G8-MRA-152 (contributed by David Walliker), 7G8-MRA-154 (contributed by Dennis E. Kyle) and 7G8-MRA-926 (contributed by Karen Hayton and Tom Wellems) parasites were obtained through BEI Resources, NIAID, NIH. Two frozen vials (termed cell bank aliquots A and B) of 7G8 parasites from Sanaria’s working cell bank (lot: SAN03-021214 from 20. February 2014) were separately thawed and cultured in human O+ erythrocytes and in presence of 10% heat-inactivated human serum in parasite culture medium according to standard procedures ^59^. To maintain synchronized parasites, 7G8 cultures were treated either with 5% sorbitol ^60^. Tight synchronization was performed by percoll-enrichment of schizont ^61^ followed by sorbitol treatment after 4 hours of cultivation. From cell bank aliquot A ring stage parasites were collected at generations 12, 15 and 24 after thawing, from cell bank aliquot B at generations 6, 13 and 19.

Selection of *var2csa* expressing parasites was performed by panning on plastic dishes coated with 50 µg/ml bovine trachea CSA (Sigma), as described previously ^62^. Subclones of Pf7G8 cell bank aliquot A were generated by limiting dilution cloning, as described previously ^63^.

### Western blot analysis

Trophozoite stage cultures were treated with 0.075% saponin in PBS to release hemoglobin from the erythrocytes. Parasites and membrane ghosts were pelleted by centrifugation, washed three times in PBS containing protease inhibitors (cOmplete EDTA free, Roche), and extracted in 2 x Laemmli buffer. The protein extracts were separated on 3–8% Tris-Acetate gels (Invitrogen) and transferred to nitrocellulose membranes (Millipore). The blots were probed with monoclonal mouse anti-ATS (6HI) antibody ^64^ or rabbit anti-*Plasmodium* aldolase antibody (abcam, ab207494) as loading control.

### gDNA purification for whole genome sequencing

For gDNA sequencing, 150 mL Pf cell culture with >10% parasitemia were harvested for the 7G8 cell bank aliquot A subclones A1G9, A2E10, and A2G2 and gDNA isolation was performed using the MasterPure™ Complete DNA purification kit (Lucigen) according to the manufacturer’s instructions “Cell samples” followed by “Complete Removal of RNA” with additional RNase I treatment. The gDNA samples were checked for degradation and RNA contamination on an agarose gel and quantified with the Qubit™ dsDNA BR Assay Kit (ThermoFischer).

### RNA purification and cDNA synthesis

Red blood cells were settled by centrifugation and completely lysed in 5 volumes of pre-warmed TRIzol (ThermoFisher). Samples were stored at -80°C until RNA purification. The RNeasy Mini kit with on-column DNase I treatment (Qiagen) was used for RNA purification. The absence of gDNA was checked for each sample using 50 ng RNA and the *skeleton-binding protein 1* (*sbp1*) primer set (Table S1). cDNA synthesis was performed as previously described ^33^.

### Quantitative real-time PCR

The LightCycler 480 (Roche) was used for quantitative real-time PCR analysis using the provided LightCycler®480 software release 1.5.1.62 SP3 as previously described ^33^. Briefly, cDNA template was mixed with QuantiTect SYBR Green PCR reagent (Qiagen) and 0.3 µM sense and antisense primer in a final volume of 10 µl. Reactions were incubated at 95°C for 15 min, then subjected to 40 cycles of 95°C for 15 s and 60°C for 1 min followed by a melting step (60–95°C). The specificity of each primer pair was confirmed by dissociation curve analysis after each qPCR run. Ct calculation was done using the fit points analysis method provided by the software. Expression of *arginyl-tRNA synthetase* (PF3D7_1218600) was used for normalization and Ct values obtained by analysis of 2.5 ng gDNA from Sanaria’s working cell bank parasites were used for calibration. Relative quantification of the 7G8 *var* repertoire by 2^-ΔΔCt^ analysis was performed using newly designed primer sets for 7G8 (Table S1). Furthermore, primer pairs targeting the housekeeping gene *fructose-bisphosphate aldolase* as well as the ring stage control *sbp1* were included (Table S1). Relative expression data (RELEXP) were corrected for amplification efficiency of each newly designed primer pair, which was determined by dilution of a single gDNA from 7G8 over 5–6 logs of concentration (Table S1). For further characterization, the full 7G8 primer set was checked for cross-reactivity with NF54 gDNA ^33^, but only primers targeting the partial gene Pf7G8_120024200 produced a specific amplicon with NF54 gDNA.

### RNA and gDNA sequencing

RNA from cell bank parasites (aliquot B, generation 13 after thawing) was purified and processed as previously described ^65^. Briefly, absence of genomic DNA was checked, human globin mRNA was depleted, and RNA quantity and quality were assessed with an Bioanalyzer (RIN value 8.4). A 100–500 bp library was prepared using the NEBNext Ultra Directional RNA Library Prep Kit for Illumina including the amplification with the KAPA polymerase, and RNA sequencing on an Illumina HiSeq4000 was conducted by BGI Genomics Co., Hongkong. Approximately 12.6 million clean reads were obtained for pre-mosquito cell bank parasites, resulting in 6.3 million paired-end 100 bp reads.

Library construction of gDNA and paired-end 150 bp sequencing of approximately 350 bp fragments (range: 230–430 bp) on the DNBseq platform was done by BGI Genomics Co., Hongkong with coverage of approximately 130x (range: 129x–136x).

### RNA-seq data analysis

After successful quality control of RNA-seq reads with FastQC v0.11.8 (http://www.bioinformatics.babraham.ac.uk/projects/fastqc/)^66^, the reads were aligned to the Pf 7G8 genome available from the PlasmoDB genome database release 45 or to the Pf 7G8 exon 1 *var* gene sequences (Data S1) ^37^ using the RNA-seq aligner STAR v2.7.3a ^67^ allowing for a maximum mismatch of 1 (--outFilterMismatchNmax 1). Prior to read alignment, STAR was used to generate an index using the genome sequence fasta file (PlasmoDB-45_Pfalciparum7G8_Genome.fasta) or the exon 1 *var* gene sequences from Data S1. The mapped reads in Sequence Alignment/Map (SAM) format were then summarized using the featureCounts ^68^ function of the Rsubread R package ^69^, filtering for a minimum fragment length of 85 bp for paired-end reads (minFragLength=85), as was done previously ^65^. While the annotation file PlasmoDB-45_Pfalciparum7G8.gff was used to summarize read counts across the entire Pf 7G8 genome, a simplified annotation format (SAF) file was built from *var* gene index files and supplied as annotation file to featureCounts.

The R package edgeR ^70^ was used to compute FPKM gene expression values using its rpkm function. The gene lengths of transcripts required for FPKM normalization were extracted from the PlasmoDB-45_Pfalciparum7G8_AnnotatedTranscripts.fasta file or the coding_nt.fa file (for the specific *var* gene assembly mapping) by indexing it using SAMtools ^71^ faidx and extracting the first two columns containing sequence name and length.

Raw and normalized RNA-seq data were submitted to the BioStudies ArrayExpress collection (E-MTAB-12157).

### gDNA-seq data analysis, mapping and variant calling

The raw reads were trimmed, mapped and variant calls were made using the CLC Genomics Workbench version 21 for Linux. Since all subclonal lines were cloned just before sequencing by limiting dilution, only variant calls with >90% reads mapped to the alternative allele were considered. Manual inspection further reduced the number of reliable variants, as the AT-rich genome of Pf is prone to sequencing errors within repetitive regions. Results and raw data from gDNA-seq were submitted to the BioStudies ArrayExpress collection (E-MTAB-12158).

### Chromatin immunoprecipitation analysis (ChIP-qPCR)

Chromatin immunoprecipitation was preformed essentially as described previously ^72^. Briefly, 1% paraformaldehyde-crosslinked chromatin was sheared by sonication and the soluble fraction was used for immunoprecipitation overnight using protein G coupled sepharose beads (GE Healthcare). Antibodies used for ChIP were rabbit anti-H3 (Abcam ab1791), anti-H3K9me3 (Active Motif 39161), anti-H3K27ac (Abcam ab4729), and anti-H2A.Z ^24^. Washed immune complexes were eluted and de-crosslinked over night at 45°C in the presence of 500 mM NaCl. After proteinase K treatment for 1h at 37°C, the DNA was purified from each ChIP and input sample using the MinElute kit (Qiagen 28006) and analyzed by qPCR on an Applied Biosystems 7900HT fast real-time PCR system using SYBR Green PCR MasterMix (ThermoFisher Scientific 4309155) and 0.9 µM sense and antisense primers (Table S1) in a final volume of 10 µl. ChIP enrichment at each genomic locus was calculated as % of input DNA, and histone modifications and H2A.Z were normalized relative to H3.

### Data analysis and statistics

Categorical variables were displayed as frequencies and percentages, and continuous variables as median and interquartile rage (IQR).

The differences between the two molecular methods qPCR and RNA-seq were analyzed using a Bland-Altman plot showing the agreement between two quantitative measurements. The differences between log-transformed measurements were plotted on the y-axis and the mean of the respective qPCR and RNA-seq results on the x-axis. Thus, the graph shows the deviation of the analysis results with respect to the expression level.

The expressions of *var* genes per patient were summarized in a heat map accompanied by a dendrogram of hierarchical cluster analysis applied to the expression pattern of patients. To correct for individual differences in the overall *var* transcript levels, the level for each *var* gene was normalized against total *var* transcript level in each sample. *Var* gene expressions within patient groups were summarized using the median and interquartile range (IQR). Boxplots showing the minimum, maximum, IQR, and median were used to graphically represent group expressions. Outliers, defined as values above or below the median +/-1.5 times the IQR, are plotted outside the whiskers of the boxplot. Correlations between individual *var* gene expressions are calculated using the Pearson correlation coefficient (PCC) with the log-transformed measurements.

## Data Availability

All data produced in the present work are contained in the manuscript or are available online at the BioStudies ArrayExpress collection (E-MTAB-12158, E-MTAB-12157).

https://www.ebi.ac.uk/biostudies/arrayexpress/studies/E-MTAB-12157

https://www.ebi.ac.uk/biostudies/arrayexpress/studies/E-MTAB-12158

## Funding

JH, ME, PGK, BM and AB received funding for the clinical trial and the *var* gene expression analysis by the Federal Ministry of Education and Research in the framework of the German Centre for Infection Research (DZIF) (TTU 03.702 Clinical Trial Platform and TTU 03.703 Clinical Research Group) (http://www.dzif.de/). This work was supported by the DFG Research Infrastructure NGS_CC (project #1016) as part of the Next Generation Sequencing Competence Network (project 423957469). NGS analyses were carried out at the production site WGGC-Bonn. AB and MP received the DFG grants BA 5213/6-1 and PE 1618/4-1 (project #433302244), respectively, as part of the DFG Sequencing call 2019. JSWM, YDH and AB were funded by the German Research Foundation (DFG) grants BA 5213/3-1 (project #323759012) and BA 5213/6-1 (project #433302244). Manufacture of Sanaria® PfSPZ Vaccine, PfSPZ Challenge (NF54) and PfSPZ Challenge (7G8) was funded in part by the National Institute of Allergy and Infectious Diseases of the National Institutes of Health under SBIR award numbers 5R44AI058375 and 5R44AI055229. JCS, KAM and AD were funded by awards U19 AI110820 and R01 AI141900, from the National Institute for Allergy and Infectious Diseases, National Institutes of Health. The funders had no role in study design, data collection and analysis, decision to publish, or preparation of the manuscript.

## Acknowledgements

We thank all volunteers who participated in the trials at the Institute of Tropical Medicine in Tübingen, Germany, and the entire study team. We thank Sanaria Inc. for providing the 7G8 working cell bank parasites and the PfSPZ Challenge (7G8) used in the CHMI trials and Balázs Horváth for his help with data analysis, mapping and variant calling. The following reagents were obtained through BEI Resources, NIAID, NIH: *Plasmodium falciparum*, Strain 7G8, MRA-152, contributed by David Walliker, MRA-154, contributed by Dennis E. Kyle and MRA-926, contributed by Karen Hayton and Tom Wellems.

## Figures

**Table 1: Overview of volunteer characteristics infected with PfSPZ. Parasite counts were determined on the day of treatment/sampling by either thick blood smear (TBS) or qPCR.**

**Figure S1: Schematics of the MAVACHE and CVac-Tü3 study designs**.

**Figure S2: Ranking of *var* genes according to transcript levels detected in samples from malaria-naïve volunteers infected with PfSPZ 7G8**. The median *var* transcript level relative to the *arginyl-tRNA synthetase* transcript level with IQR is shown for 11 volunteer samples. Group affiliation of *var* genes is indicated by the color code: A-type *var* genes in red, the subfamily *var1* in dark red, B-type genes in blue, group C genes in green, and the *var2csa* gene (group E) in yellow.

**Figure S3: *Var* gene expression profiles of 7G8 cell bank parasites derived from Sanaria (aliquot A and B) and BEI Resources (MRA-152, MRA-926, MRA-154). (A**) Heat map showing *var* expression of 7G8 parasites from Sanaria cell bank aliquots A and B and of three 7G8 aliquots deposited at BEI resources by different providers. Expression of each *var* gene is normalized to expression of *arginyl-tRNA synthetase*. (**B**) Pie charts showing the proportion of *var* gene expression by group for the different 7G8 lines. The names of *var* genes are indicated, and *var* gene groups are colored according to the scheme: A in red, A-*var1* in dark red, B in blue and C in green.

**Figure S4: *Var* gene expression profiles of CSA-selected 7G8 cell bank parasites after cultivation for up to 100 replication cycles after selection**. Heat map showing the *var* expression of 7G8 cell bank parasites (aliquot A) selected on CSA to express *var2csa* after cultivation for up to 100 parasite replications. The expression of each *var* gene is normalized against the expression of *arginyl-tRNA synthetase*. The names of *var* genes are indicated, and *var* gene groups are colored according to the scheme: A in red, A-*var1* in dark red, B in blue and C in green.

**Figure S5: qPCR on gDNA from 7G8 subclones to confirm presence of each *var* gene variant in the different genomes**. Shown are raw Ct values from about 2.5 ng gDNA used as template per qPCR reaction. The Ct values from the cell bank aliquot A bulk culture are marked in black for reference, and a line is drawn at the mean.

**Table S1: The 7G8 *var* gene repertoire and the domain composition of the encoding PfEMP1 variants according to the classification from Rask *et al***. ^5^. Question marks indicate unknown features of the *var* gene/PfEMP1. Gene IDs and gene type annotation according to PlasmoDB Release 58 (23. June 2022). ATS: Acidic terminal segment; CIDR: Cysteine-rich interdomain region; DBL: Duffy-binding like domain; NTS: N-Terminal segment; UPS: upstream sequence

**Table S2: Oligonucleotides used in this study**.

**Table S3: qPCR data on 7G8 *var* gene expression in pre-mosquito cell bank parasites (A) and in parasites from malaria-naïve volunteers (B)**.

**Table S4: *Var* gene expression in MRA-152**, 7G8 **pre-mosquito cell bank parasites (aliquot A) and 7G8 subclones before and after CSA selection**.

**Table S5: Unique genomic variants of 7G8 subclones A1G9 (var2csa-expressing subclone, control) versus A2G2 and A2E10 (Pf7G8_040025600-expressing subclones) determined by gDNA-seq**.

**Table S6: ChIP-qPCR and *var* expression profiles from 7G8 pre-mosquito cell bank parasites (aliquot A) and 7G8 subclones before and after CSA selection**.

**Data S1: All *var* exon 1 sequences used for design of the oligonucleotides and RNA-seq mapping**

(coding_nt.fa) ^37^.

## Notes

### Competing Interest Statement

The authors have declared no competing interest.

### Author Declarations

The ethics committee of the University Clinic and the Medical Faculty of the University of Tuebingen approved both studies, MAVACHE and CVac-Tue3, of which samples were examines in this work, and the U.S. Food and Drug Administration Agency (FDA) provided regulatory oversight.

