## Supplementary Figures for "The exception that proves the rule: Virulence gene expression at the onset of *Plasmodium falciparum* blood stage infections"

**MAVACHE**

Sequential optimization of dose and schedule of PfSPZ Vaccine, verified by randomized, controlled, double-blind immunization and controlled human malaria infection in malaria-naïve, healthy adult volunteers in Germany

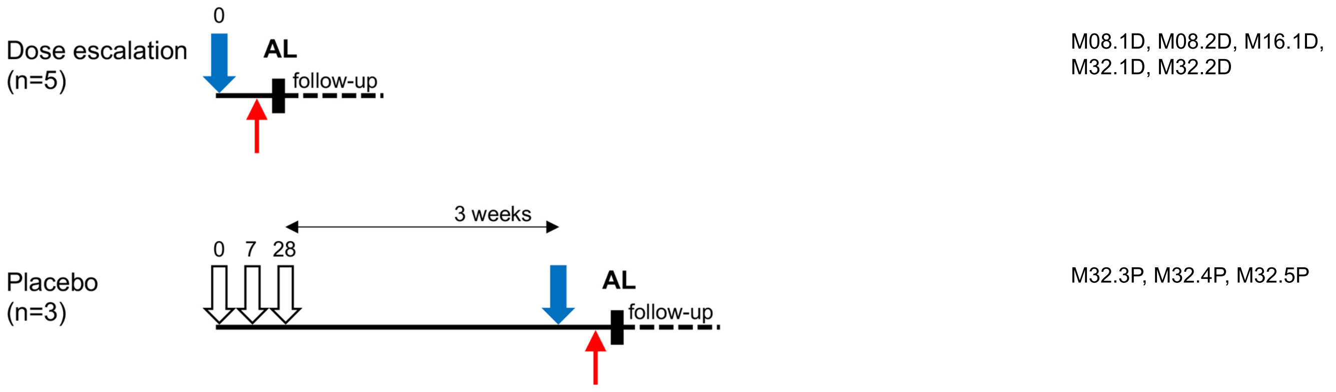


**CVac-Tü3**

Safety and protective efficacy of a simplified *P. falciparum* sporozoite Chemoprophylaxis Vaccine (PfSPZ-CVac) regimen in healthy malaria-naïve adults in Germany

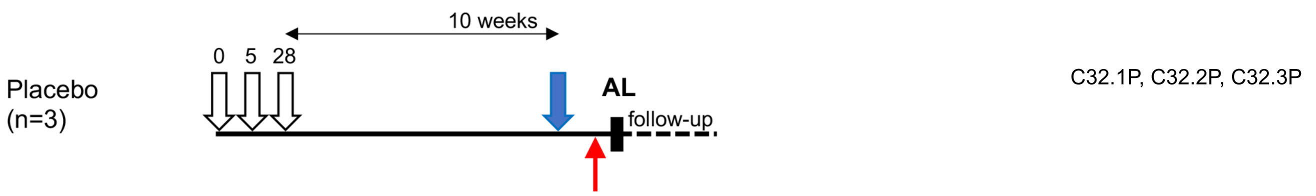


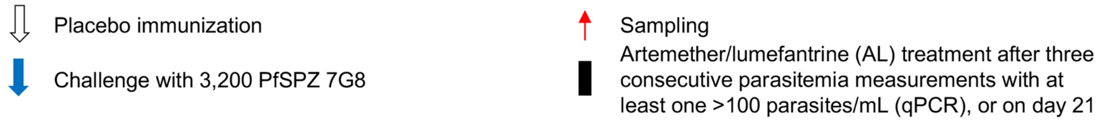

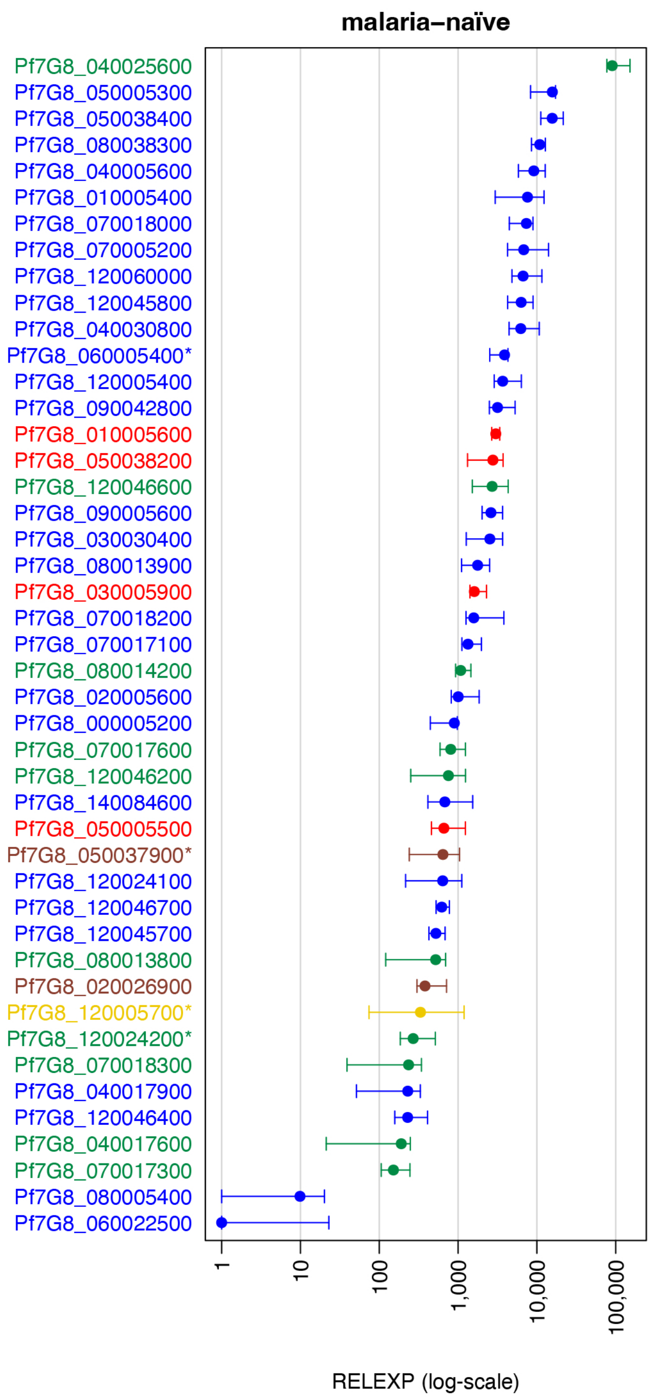
malaria–naïve
Pf7G8_040025600
Pf7G8_050005300
Pf7G8_050038400
Pf7G8_080038300
Pf7G8_040005600
Pf7G8_010005400
Pf7G8_070018000
Pf7G8_070005200
Pf7G8_120060000
Pf7G8_120045800
Pf7G8_040030800
Pf7G8_060005400*
Pf7G8_120005400
Pf7G8_090042800
Pf7G8_010005600
Pf7G8_050038200
Pf7G8_120046600
Pf7G8_090005600
Pf7G8_030030400
Pf7G8_080013900
Pf7G8_030005900
Pf7G8_070018200
Pf7G8_070017100
Pf7G8_080014200
Pf7G8_020005600
Pf7G8_000005200
Pf7G8_070017600
Pf7G8_120046200
Pf7G8_140084600
Pf7G8_050005500
Pf7G8_050037900*
Pf7G8_120024100
Pf7G8_120046700
Pf7G8_120045700
Pf7G8_080013800
Pf7G8_020026900
Pf7G8_120005700*
Pf7G8_120024200*
Pf7G8_070018300
Pf7G8_040017900
Pf7G8_120046400
Pf7G8_040017600
Pf7G8_070017300
Pf7G8_080005400
Pf7G8_060022500
1
10
100
1,000
10,000
100,000
RELEXP (log-scale)

Figure S3

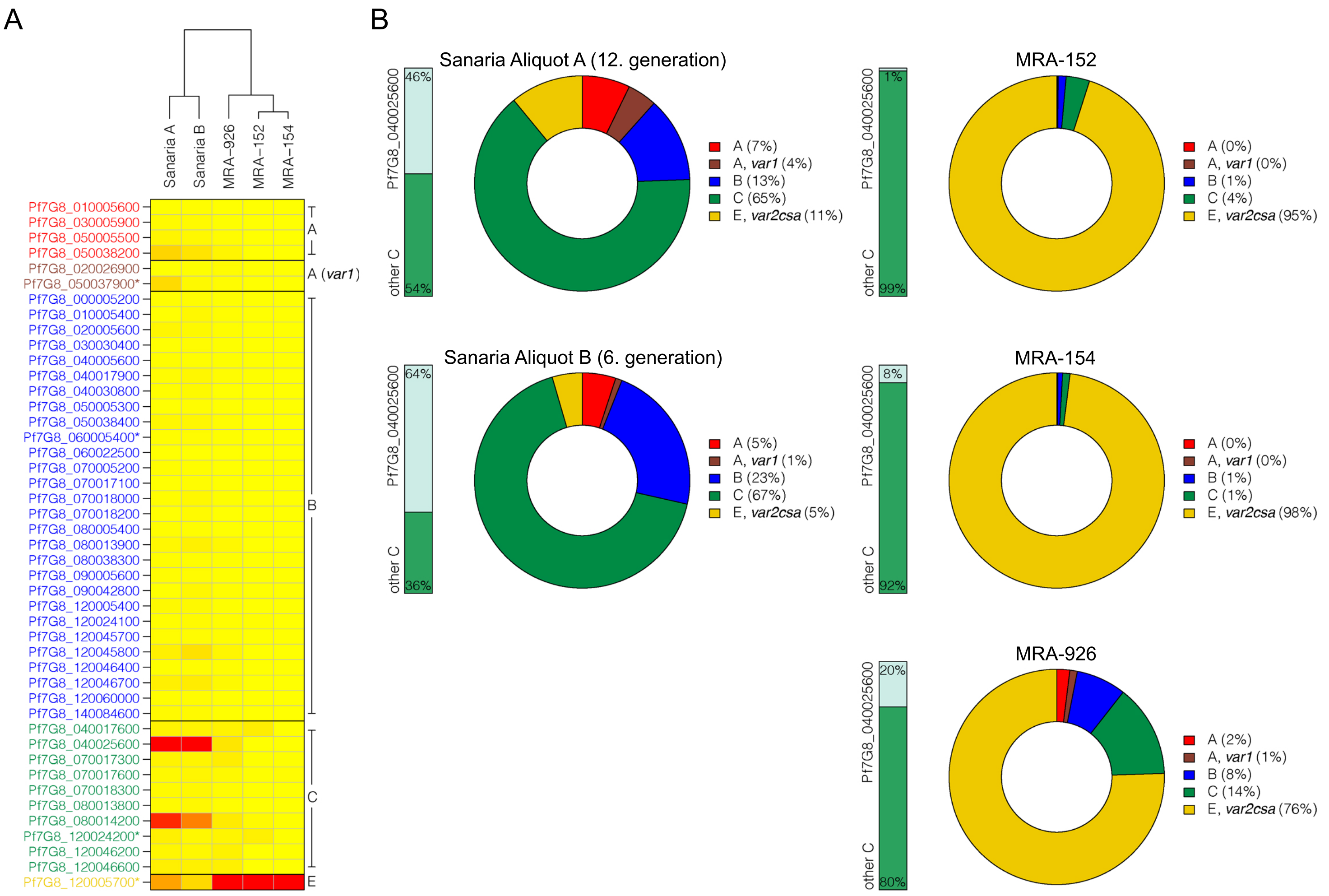

A
Sanaria A
Sanaria B
MRA-926
MRA-152
MRA-154
Pf7G8_010005600
Pf7G8_030005900
Pf7G8_050005500
Pf7G8_050038200
Pf7G8_020026900
Pf7G8_050037900*
Pf7G8_000005200
Pf7G8_010005400
Pf7G8_020005600
Pf7G8_030030400
Pf7G8_040005600
Pf7G8_040017900
Pf7G8_040030800
Pf7G8_050005300
Pf7G8_050038400
Pf7G8_060005400*
Pf7G8_060022500
Pf7G8_070005200
Pf7G8_070017100
Pf7G8_070018000
Pf7G8_070018200
Pf7G8_080005400
Pf7G8_080013900
Pf7G8_080038300
Pf7G8_090005600
Pf7G8_090042800
Pf7G8_120005400
Pf7G8_120024100
Pf7G8_120045700
Pf7G8_120045800
Pf7G8_120046400
Pf7G8_120046700
Pf7G8_120060000
Pf7G8_140084600
Pf7G8_040017600
Pf7G8_040025600
Pf7G8_070017300
Pf7G8_070017600
Pf7G8_070018300
Pf7G8_080013800
Pf7G8_080014200
Pf7G8_120024200*
Pf7G8_120046200
Pf7G8_120046600
Pf7G8_120005700*
A
A (var1)
B
C
E
B
Sanaria Aliquot A (12. generation)
Pf7G8_040025600
other C
46%
54%
A (7%)
A, var1 (4%)
B (13%)
C (65%)
E, var2csa (11%)
MRA-152
1%
99%
A (0%)
A, var1 (0%)
B (1%)
C (4%)
E, var2csa (95%)
Sanaria Aliquot B (6. generation)
64%
36%
A (5%)
A, var1 (1%)
B (23%)
C (67%)
E, var2csa (5%)
MRA-154
8%
92%
A (0%)
A, var1 (0%)
B (1%)
C (1%)
E, var2csa (98%)
MRA-926
20%
80%
A (2%)
A, var1 (1%)
B (8%)
C (14%)
E, var2csa (76%)

### Figure S4

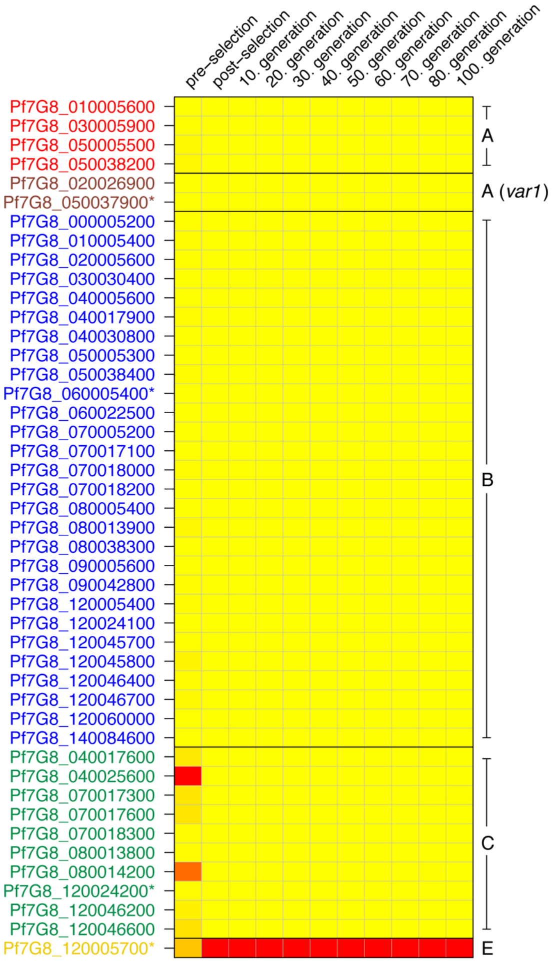

pre-selection
post-selection
10. generation
20. generation
30. generation
40. generation
50. generation
60. generation
70. generation
80. generation
100. generation
Pf7G8_010005600
Pf7G8_030005900
Pf7G8_050005500
Pf7G8_050038200
Pf7G8_020026900
Pf7G8_050037900*
Pf7G8_000005200
Pf7G8_010005400
Pf7G8_020005600
Pf7G8_030030400
Pf7G8_040005600
Pf7G8_040017900
Pf7G8_040030800
Pf7G8_050005300
Pf7G8_050038400
Pf7G8_060005400*
Pf7G8_060022500
Pf7G8_070005200
Pf7G8_070017100
Pf7G8_070018000
Pf7G8_070018200
Pf7G8_080005400
Pf7G8_080013900
Pf7G8_080038300
Pf7G8_090005600
Pf7G8_090042800
Pf7G8_120005400
Pf7G8_120024100
Pf7G8_120045700
Pf7G8_120045800
Pf7G8_120046400
Pf7G8_120046700
Pf7G8_120060000
Pf7G8_140084600
Pf7G8_040017600
Pf7G8_040025600
Pf7G8_070017300
Pf7G8_070017600
Pf7G8_070018300
Pf7G8_080013800
Pf7G8_080014200
Pf7G8_120024200*
Pf7G8_120046200
Pf7G8_120046600
Pf7G8_120005700*
A
A (var1)
B
C
E

Figure S5

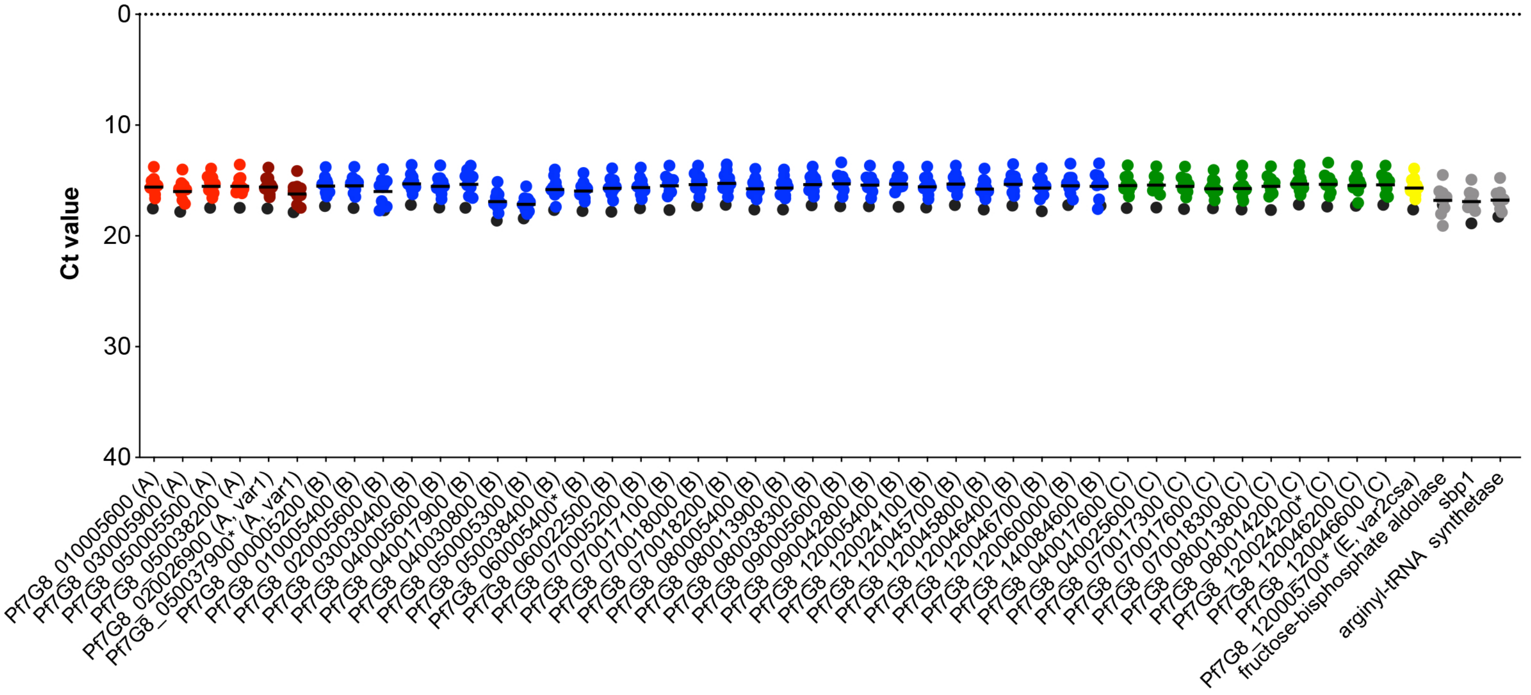
Ct value
0
10
20
30
40
Pf7G8_010005600 (A)
Pf7G8_030005900 (A)
Pf7G8_050005500 (A)
Pf7G8_050038200 (A)
Pf7G8_020026900 (A, var1)
Pf7G8_050037900* (A, var1)
Pf7G8_000005200 (B)
Pf7G8_010005400 (B)
Pf7G8_020005600 (B)
Pf7G8_030030400 (B)
Pf7G8_040005600 (B)
Pf7G8_040017900 (B)
Pf7G8_040030800 (B)
Pf7G8_050005300 (B)
Pf7G8_050038400 (B)
Pf7G8_060005400* (B)
Pf7G8_060022500 (B)
Pf7G8_070005200 (B)
Pf7G8_070017100 (B)
Pf7G8_070018000 (B)
Pf7G8_070018200 (B)
Pf7G8_080005400 (B)
Pf7G8_080013900 (B)
Pf7G8_080038300 (B)
Pf7G8_090005600 (B)
Pf7G8_090042800 (B)
Pf7G8_120005400 (B)
Pf7G8_120024100 (B)
Pf7G8_120045700 (B)
Pf7G8_120045800 (B)
Pf7G8_120046400 (B)
Pf7G8_120046700 (B)
Pf7G8_120060000 (B)
Pf7G8_140084600 (B)
Pf7G8_040017600 (C)
Pf7G8_040025600 (C)
Pf7G8_070017300 (C)
Pf7G8_070017600 (C)
Pf7G8_070018300 (C)
Pf7G8_080013800 (C)
Pf7G8_080014200 (C)
Pf7G8_120024200* (C)
Pf7G8_120046200 (C)
Pf7G8_120046600 (C)
Pf7G8_120005700* (E, var2csa)
fructose-bisphosphate aldolase
sbp1
arginyl-tRNA synthetase
